# COVID19 Diagnosis Using Chest X-rays and Transfer Learning

**DOI:** 10.1101/2022.10.09.22280877

**Authors:** Jonathan Stubblefield, Jason Causey, Dakota Dale, Jake Qualls, Emily Bellis, Jennifer Fowler, Karl Walker, Xiuzhen Huang

**Affiliations:** Arkansas AI-Campus, Molecular Biosciences, Arkansas State University, Jonesboro, AR, USA; Arkansas AI-Campus, Computer Science, Arkansas State University, Jonesboro, AR, USA; Arkansas AI-Campus, Computer Science, University of Arkansas, at Fayetteville, AR, USA; Arkansas AI-Campus, Computer Science & Math, University of Arkansas at, Pine Bluff, AR, USA; Arkansas AI-Campus, Arkansas State University, Dept. of Computational, Biomedicine, Cedars Sinai, Medical Center

**Keywords:** COVID19, Chest X-Ray, Medical Imaging, Machine Learning, Transfer Learning

## Abstract

A pandemic of respiratory illnesses from a novel coronavirus known as Sars-CoV-2 has swept across the globe since December of 2019. This is calling upon the research community including medical imaging to provide effective tools for use in combating this virus. Research in biomedical imaging of viral patients is already very active with machine learning models being created for diagnosing Sars-CoV-2 infections in patients using CT scans and chest x-rays. We aim to build upon this research. Here we used a transfer-learning approach to develop models capable of diagnosing COVID19 from chest x-ray. For this work we compiled a dataset of 112120 negative images from the Chest X-Ray 14 and 2725 positive images from public repositories. We tested multiple models, including logistic regression and random forest and XGBoost with and without principal components analysis, using five-fold cross-validation to evaluate recall, precision, and f1-score. These models were compared to a pre-trained deep-learning model for evaluating chest x-rays called COVID-Net. Our best model was XGBoost with principal components with a recall, precision, and f1-score of 0.692, 0.960, 0.804 respectively. This model greatly outperformed COVID-Net which scored 0.987, 0.025, 0.048. This model, with its high precision and reasonable sensitivity, would be most useful as “rule-in” test for COVID19. Though it outperforms some chemical assays in sensitivity, this model should be studied in patients who would not ordinarily receive a chest x-ray before being used for screening.

**CCS CONCEPTS:**

- Life and Medical Sciences • Machine Learning • Artificial Intelligence

**Reference format:** Jonathan Stubblefield, Jason Causey, Dakota Dale, Jake Qualls, Emily Bellis, Jennifer Fowler, Karl Walker and Xiuzhen Huang. 2022. COVID19 Diagnosis Using Chest X-Rays and Transfer Learning.

## 1 BACKGROUND

Beginning in December of 2019, a novel, Human coronavirus emerged in Wuhan, China [4]. Though similar to the previous SARS and MERS viruses, this virus proved to be more infectious and quickly became a global pandemic [4]. Causing pneumonia-like symptoms, a total of 2,552,687 deaths have been attributed to this virus as of March 3, 2021 [4, 12]. The elderly are especially vulnerable to this virus [8].

Routine testing for this virus usually involves a nasopharyngeal or oropharyngeal swab [13]. The sample is then sent to an outside facility for determination of viral load by polymerase chain reaction (PCR) [13]. Some rapid tests exist but are not used as prevalently [13]. This test is time consuming, taking several days for results to be returned. It also expends valuable reagents and testing kits that are in limited supply. A faster, less expensive modality for diagnosing the novel coronavirus (COVID19) would be valuable in hospital settings when patients are too sick to wait for answers. Rapid chemical testing for COVID19 has been made available but suffers from the same problem of consuming reagents and test kits [16]. Additionally, rapid antigen tests have less sensitive compared to other tests [16].

Other researchers have been explored the marks left by COVID19 on radiographic images of the patient’s lungs. COVID19 presents radiologically like an atypical or organizing pneumonia, but also has some distinctive characteristics [12]. On plain-film x-ray, patchy or diffuse opacities with the texture of consolidation or ground-glass may be visible, but the unique imaging features of COVID19 are much more apparent in a CT scan [12]. CT scans of COVID19 infected patients show ground-glass opacities, crazy paving, airspace consolidation, bronchovascular thickening, and traction bronchiectasis [12].

Given these distinctive findings, machine learning algorithms have been developed to diagnose cases of the virus with CT data [9]. A meta-analysis of machine learning models used for diagnosis of COVID19 based on medical imaging found 18 papers applying deep-learning to CT scans. These deep-learning models performed well on their respective datasets with reported area-under the receiver operating characteristic curve (AUROC) between 0.7 and 1.0 [17].

However, CT scans come with their own problems. Compared to a plain-film x-ray, CT scans use much larger amounts of radiation, presenting higher risk to the patient. For this reason, physicians use them sparingly. A diagnostic algorithm for COVID19 infection using a plain-film chest x-ray as its input would greatly economize the patient’s radiation exposure. The same meta-analysis found 22 papers using deep-learning to analyze plain-film chest x-rays [16]. These models also exhibited good performance on their datasets with accuracies ranging from 0.88 to 0.99 [17].

Our goal is to improve upon the existing technology for deep-learning diagnosis of COVID19 using chest x-rays by including larger number of images in the training set and incorporating a transfer learning approach. We will use a pre-trained CheXNet model to extract imaging features from chest x-rays for use in a downstream. This approach was used by this research group previously for predicting the etiology of acute shortness of breath in an ER setting [15].

### Summary of this work

For this project, we used a transfer-learning approach to develop a model capable of diagnosing COVID19 from chest x-ray. For this project we compiled a dataset of 112120 negative images from the Chest X-Ray 14 and 2725 positive images from public repositories [5, 6, 7, 10, 14, 18. 19, 20]. Features were extracted from the images using a CheXNet trained on Chest X-Ray 14 [1, 5]. The output layer and penultimate layer were used, giving a total of 1038 raw features. The feature data was split into five folds for cross-validation. Multiple downstream models, including logistic regression and random forest and XGBoost with and without principal components analysis, were tested using cross-validation to evaluate recall, precision, and f1-score [2, 23]. These models were compared to a pre-trained deep-learning model for evaluating chest x-rays called COVID-Net [3]. Our best model was XGBoost with principal components with a recall, precision, and f1-score of 0.692, 0.960, 0.804 respectively [2]. This model greatly outperformed COVID-Net which scored 0.987, 0.025, 0.048 [3]. This drastic improvement in performance is most likely due to the expanded dataset used in the training of our model compared to the dataset used to train COVID-Net [3]. This model, with its high precision and reasonable sensitivity, would be most useful as “rule-in” test for COVID19. Though it outperforms some chemical assays in sensitivity, this model should be studied in patients who would not ordinarily receive a chest x-ray before being used for screening [16].

## 2 METHODS

### 2.1 Comparison Model

For the purposes of comparison, we chose the COVIDNet-CXR4-A release of COVIDNet [3]. The model reports an accuracy of 0.943 and sensitivity of 0.950 on its own dataset [3]. For an accurate comparison, this model will be re-evaluated on the test-folds used to evaluate the rest of the models.

### 2.2 Dataset

For this project, we created a simpler model for predicting the presence or absence of COVID19 in patients with symptoms of pneumonia. To this end, we used a dataset containing a mixture of positive and negative chest x-rays. Conveniently, the Chest X-Ray 14 dataset was published prior to the outbreak of COVID19, and all images contained within are negative by default [5]. We obtained positively labeled cases of COVID19 by using the public repositories referenced in the COVIDNet Github [6, 7, 10, 14]. Additional positive cases were obtained from the cancer imaging archive [18, 19, 20]. The dataset was split into 5 folds for cross-validation. In total, there were 112,120 negative images and 2,725 positive images.

### 2.3 Features and Principal Components

We used a transfer learning approach with the previously trained CheXNet model [1]. CheXNet was trained on the Chest X-Ray 14 dataset to predict the presence or absence of 14 different medical anomalies including atelectasis, cardiomegaly, effusion, infiltration, mass, nodule, pneumonia, pneumothorax, consolidation, edema, emphysema, fibrosis, pleural thickening, and hernia [1, 5]. We will use the outputs of CheXNet as a feature layer describing each image. Additionally, we will expand this dataset with an additional 1024 features from the layer immediately before the final classification layer. Though this expanded set of features will provide a richer dataset with which to work, it will also add a great deal to the computational complexity. It is unlikely that all obtained features will contribute to the ability of the model to discriminate between COVID19 infection and other chest x-rays. To reduce the expanded set of features to a more manageable size, we used principal component analysis to reduce the total number of features with automatic choice of dimensionality using Minka’s MLE [22].

### 2.4 Data Augmentation

The dataset was markedly imbalanced by class with a large number of negative images compared to positive. Imbalanced datasets present a problem for machine learning algorithms and tend to skew the trained model toward predicting the overrepresented class more often. To overcome this issue, we used a method similar SMOTE [21]. New positive cases were generated from existing positive cases by randomly selecting 2-4 positive cases and calculating the centroid of each of the extracted features for the group.

### 2.5 Logistic Regression and Trees

To analyze the features from CheXNet, we used a variety of models, including logistic regression, random forest, XGBoost, and a neural network. We performed this analysis using python and the scikit-learn and XGBoost packages for python [2, 23]. CheXNet features were obtained from all images and saved prior to training. The random forest and XGBoost were evaluated on both the raw CheXNet features and on the principal components derived from the CheXNet features. All models were evaluated using recall, precision, and f1-score.

## 3 RESULTS

We conducted the evaluation of the models, including COVID-Net, Logistic Regression, Random Forest, XGBoost, Random Forest with Principal Components, and XGBoost with Principal Components. We used the 5-fold cross validation. We reported a summary of the testing results in Table 1, with the Recall, Precision and F1_Score for each model on the same testing dataset.

**Table 1:** The average model performance on 5-fold cross-validation.

| Model | Recall | Precision | F1_Score |
| --- | --- | --- | --- |
| COVID-Net | 0.987 | 0.025 | 0.048 |
| Logistic Regression | 0.575 | 0.065 | 0.117 |
| Random Forest | 0.590 | 0.797 | 0.687 |
| XGBoost | 0.708 | 0.890 | 0.788 |
| RandomForest<br>w/Principal<br>Components | 0.505 | 0.865 | 0.637 |
| XGBoost<br>w/Principal<br>Components | 0.692 | 0.960 | 0.804 |

From the Table 1 testing results, we can draw some conclusions of the performance of the models. XGBoost had better performance than random forest. This is likely due to this method of using gradients and loss calculations to expand the decision forest, enabling greater flexibility and a closer fit with increased risk of overfitting. It is interesting to note that principal component analysis improved the performance of XGBoost while hindering the performance of random forest. It is also interesting that principal component analysis improved precision while decreasing sensitivity in both cases. The aggregate tree methods proved to be superior on this problem to logistic regression, likely owing to the greater flexibility of these non-parametric models.

Compared to COVIDNet, all attempted models performed better. COVIDNet tended to have a high sensitivity but committed several errors predicting COVID19 infection in images that were negative, as shown by its precision and f1-score. Given our dataset, all of these false-positives took place on the Chest X-Ray 14 dataset, which was not in the training set for COVIDNet. From this, we can conclude that the larger dataset for these models was a major source of performance improvement.

It is likely that XGBoost with principal component analysis could be used as a diagnostic test by physicians. This test has high precision compared to sensitivity, so it would be most useful as a “rule-in” test to confirm the presence of COVID19 infection in the lungs if it is already suspected, but the test still has a relatively high sensitivity. When comparing our model’s performance on this dataset to the characterizations of molecular tests by Bisoffi et al, our model outperforms the tested serologic and ELISA tests in sensitivity as well as two of the RT-PCR tests [16]. However, all patients in this dataset received chest x-rays, indicating the presence of symptoms that prompted physicians to obtain chest x-rays. A new clinical trial examining the performance of this model compared to a gold-standard RT-PCR on patients who would not ordinarily receive a chest x-ray would be needed before the model could be confidently used to screen asymptomatic patients for COVID19.

## 4 DISCUSSIONS OF FALSE POSITIVE IMAGES

This section we provide some discussions with our analysis of the false positive images. This may shed some lights on further research to figure out what features does the models learn from COVID-19 chest x-ray images.

### CheXNet Dataset Analysis

Only 592 COVID-19 false positive images (total 112120 images) were found when inferenced by COVID-Net. This proved that the model must learned some distinct features in COVID-19 images that can clearly separate COVID-19 images from the rest of the lung diseases. **Figure 3** showed that the percentage of each disease labels of false positive images.

**Figure 1:**
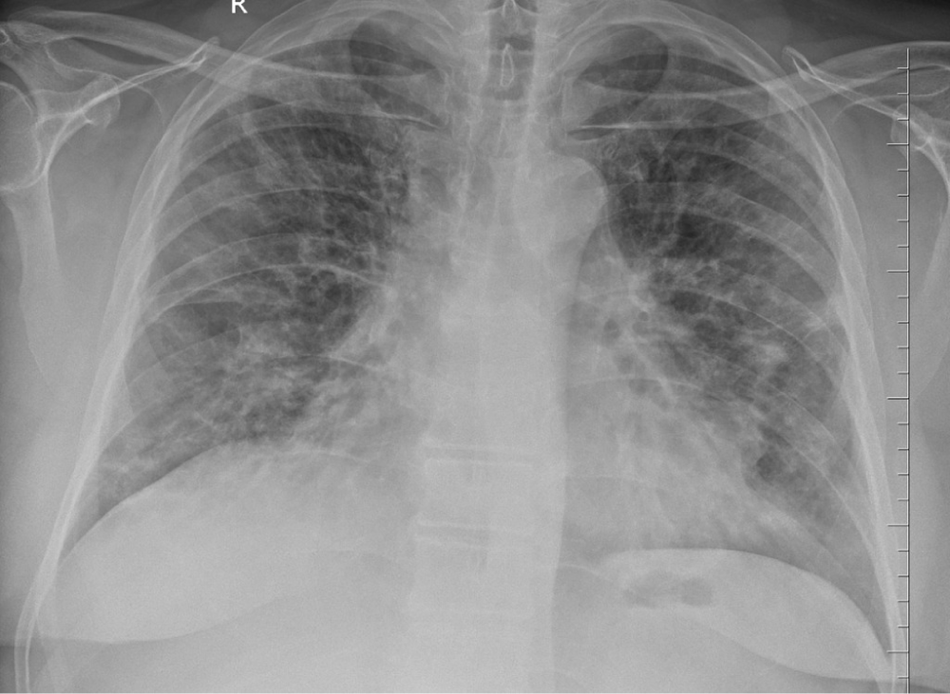
COVID19 appearance on chest x-ray. Image shows patchy bilateral opacities with diffuse ground glass appearance. Image credit: Dr. Subhan Iqbal (Radiopaedia Contributor) https://radiopaedia.org/cases/covid-19-pneumonia-101?lang=us

**Figure 2:**
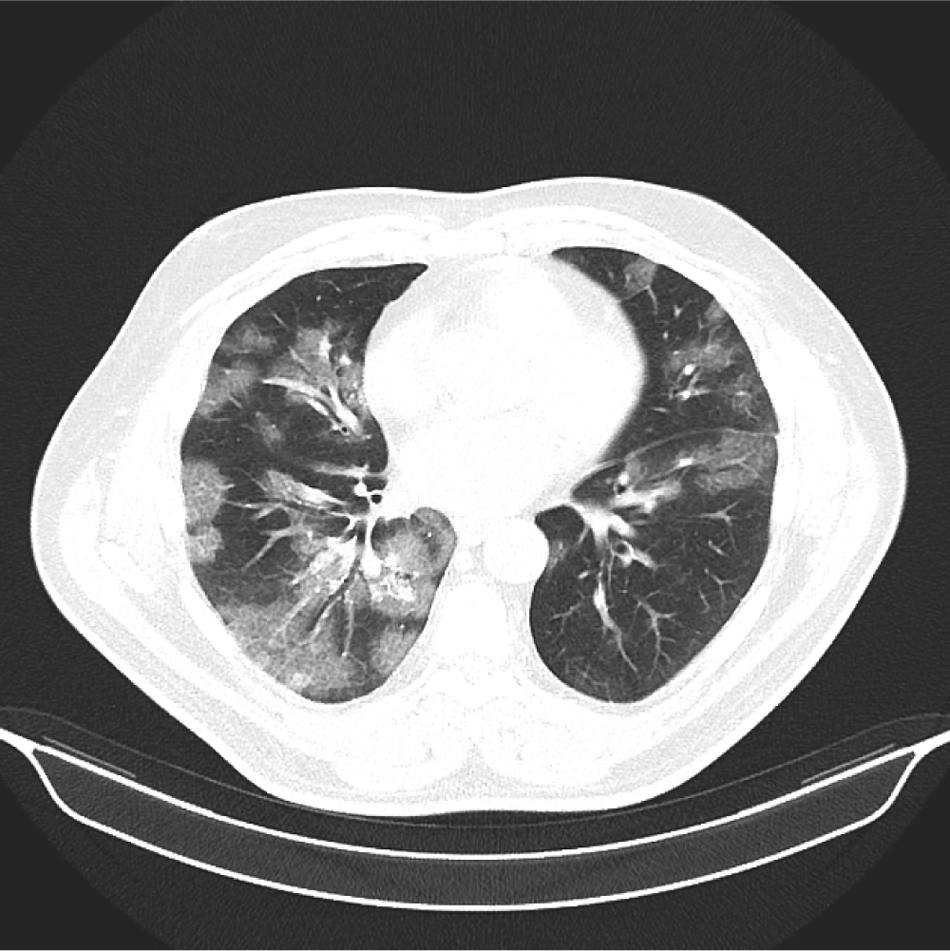
COVID19 appearance on CT scan. Image shows patchy peripheral ground glass. Image Credit: Dr Elshan Abdullayev (Radiopaedia Contributor) https://radiopaedia.org/cases/covid-19-pneumonia-45?lang=us

**Figure 3:**
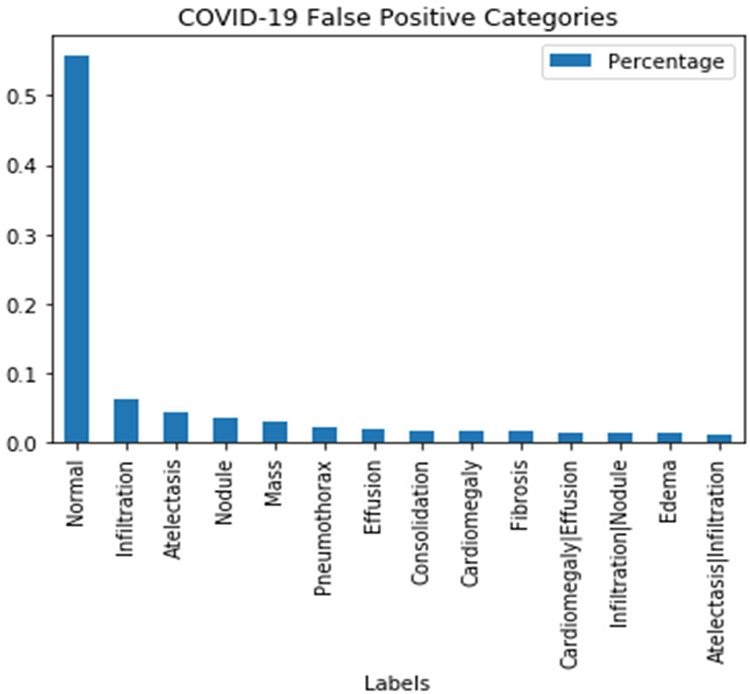
The percentage of each of the 14 disease labels among false positive images.

By going through the false positive images from different disease categories, we found that the image with no useful information can also give a high COVID-19 score (**Figure 4**). This showed that the model may learn some noise that are unique in COVID-19 images. This can be a potential reason for its good performance in distinguishing between normal and COVID-19 images.

**Figure 4:**
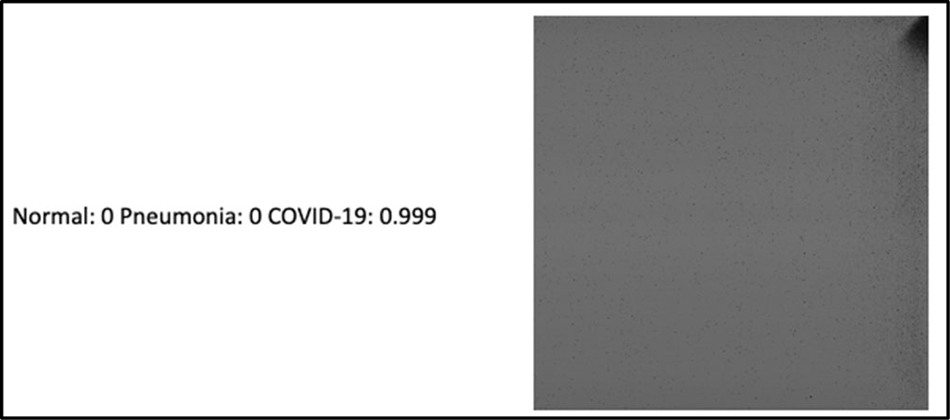
This image with no useful information can also give a high COVID-19 score.

Therefore, we also tried to inference some images that are modified artificially to check features that model learned.

### Artificially modified images

We chose an image that is in normal category and made some modifications. Please refer to **Figure 5**. By changing the original image into mosaic style (third image) or designing an image from random white and black pixels (fourth image) can give a high COVID-19 score. This showed that the model may have learned that discontinuity between black and white pixels is associated with COVID-19. Increasing intensity to the original images (second image) did not give a big change in scores.

**Figure 5:**
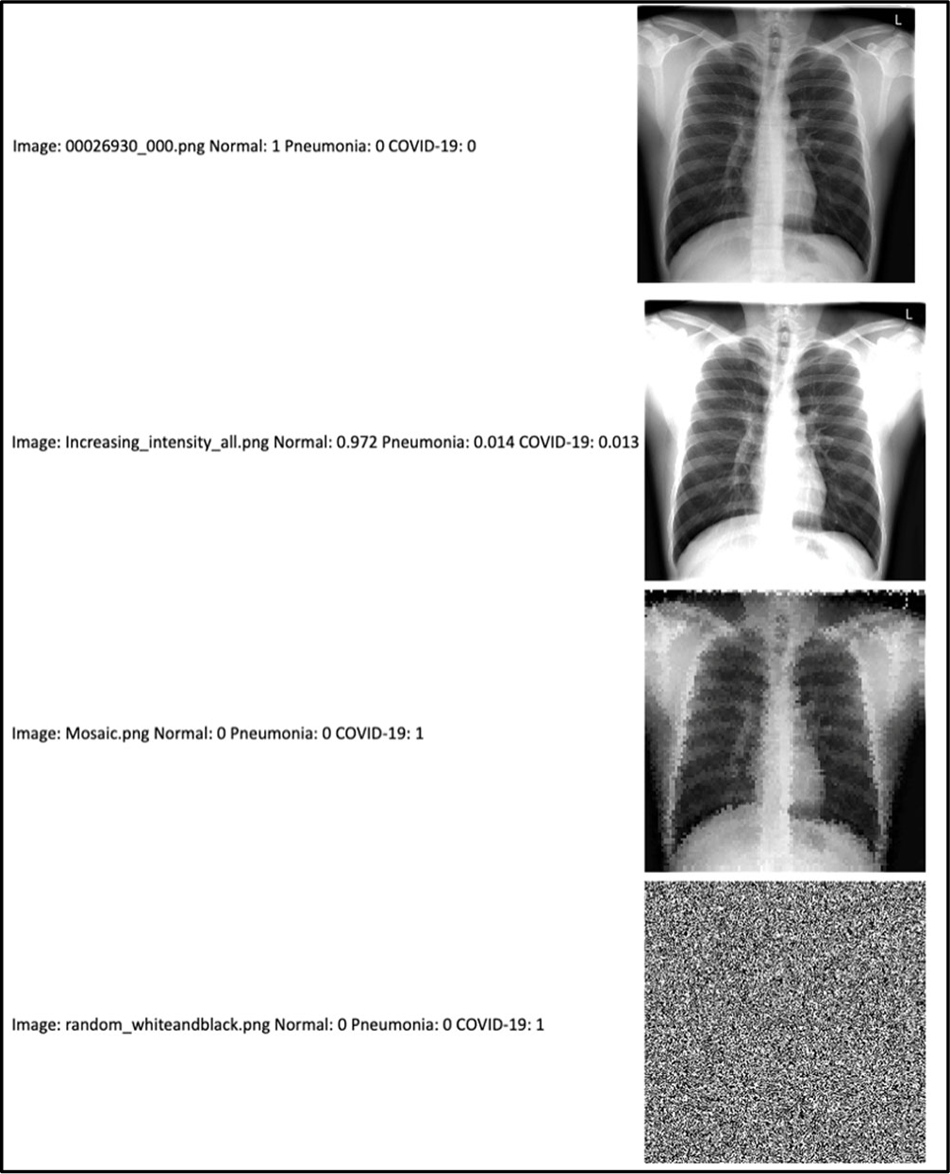
Artificially modified images. We chose an image that is in normal category and made some modifications. By changing the original image (first image) into mosaic style (third image) or designing an image from random white and black pixels (fourth image) can give a high COVID-19 score. Increasing intensity to the original images (second image) did not give a big change in scores.

### Smoothing the image

We also tried to make some modifications to COVID-19 false positive image to reverse it back to normal. Please refer to **Figure 6**. The image on the lower left is COVID-19 false positive and we added filters to smooth the images. The COVID-19 scores were dropped. This indicated that discontinuity between black and white pixels can be one of the potential features that the model learned to distinguish between Normal and COVID-19 images.

**Figure 6:**
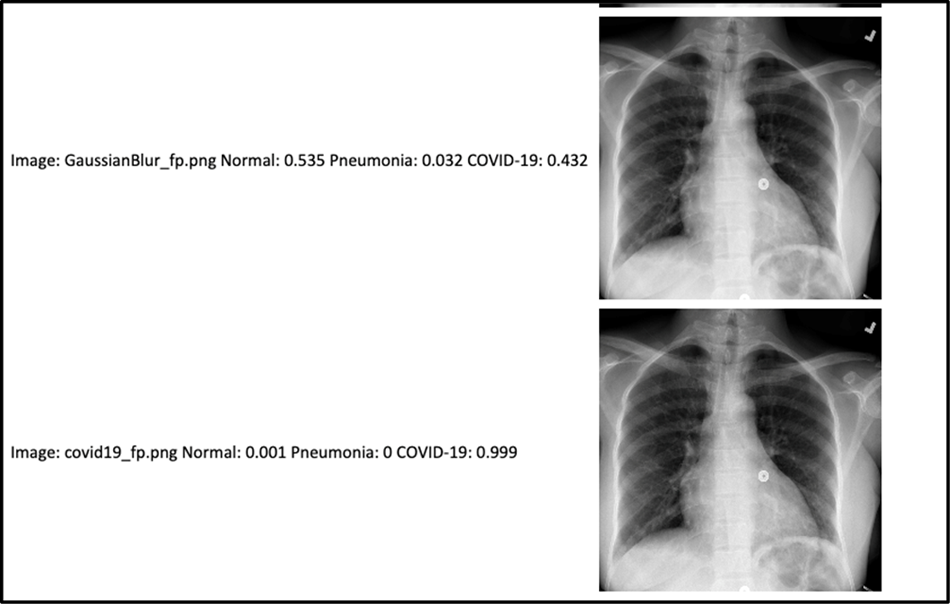
Smoothing the images. The image (bottom image) is COVID-19 false positive and we added filters to smooth the image (top image). The COVID-19 scores were dropped.

## 5 FUTURE WORK

The currently used tests for Sars-CoV-2 infection rely on chemical methods using swabs of the airway and may require sending the samples to laboratory. A computer-aided diagnostic test based on chest x-rays would not have this problem, and results could be expected quickly. Furthermore, success in distinguishing Sars-CoV-2 infection from other forms of pneumonia would also be an important step differentiating between bacterial and viral pneumonias. This critical distinction could help clinicians decide if antibiotic therapy is appropriate in their patients and improve antibiotic stewardship.

In the future, we will explore adding clinical information to the imaging information to improve model performance. To accomplish this, we are partnering with a local residency program to acquire COVID19 cases with imaging and clinical information from an established hospital system. After collecting data, we will develop a model that takes advantage of both imaging and clinical data for COVID19 diagnosis.

Additionally, this group participated this year the COVID19 EHR Dream Challenge competition [24]. This competition has proposed multiple research questions of clinical import and challenged teams from around the world to build machine learning models capable of solving them from data that could be found in a patient’s electronic health record [24]. This competition has spurred research into the diagnosis of COVID19 as well as the estimation of patient outcomes. Analysis of the resulting models will provide further insight into which elements of a patient’s electronic health record are important for diagnosis and outcomes.

## Data Availability

All data produced in the present study are available upon reasonable request to the authors.

## ACKNOWLEDGMENTS

Special thanks to the Arkansas AI-Campus coaches and participating students. This research work was partially supported by the National Science Foundation EPSCOR DART grant, and the National Science Foundation with grant number 1452211, 1553680, and 1723529, National Institute of Health grant R01LM012601, as well as was partially supported by National Institute of Health grant from the National Institute of General Medical Sciences (P20GM103429).

## Notes

### Competing Interest Statement

The authors have declared no competing interest.

### Summary of Updates

Minor edit to background section.

